# Frequent testing and immunity-based staffing will help mitigate outbreaks in nursing home settings

**DOI:** 10.1101/2020.11.04.20224758

**Authors:** Inga Holmdahl, Rebecca Kahn, James Hay, Caroline O. Buckee, Michael Mina

**Affiliations:** Center for Communicable Disease Dynamics, Department of Epidemiology, Harvard T.H. Chan School of Public Health, Boston, Massachusetts; Department of Immunology and Infectious Diseases, Harvard T.H. Chan School of Public Health, Boston, Massachusetts; Department of Pathology, Brigham and Women’s Hospital, Harvard Medical School, Boston, Massachusetts

## Abstract

**Background:** Nursing homes and other long term care facilities have been disproportionately impacted by the COVID-19 pandemic. Strategies are urgently needed to reduce transmission in these vulnerable populations. We aim to evaluate the reduction in transmission in nursing homes achieved through contact-targeted interventions and testing.

**Methods:** We developed an agent-based Susceptible–Exposed– Infectious(Asymptomatic/Symptomatic)–Recovered (SEIR) model to examine SARS-CoV-2 transmission in nursing homes. Residents and staff are modelled individually; residents are split into two cohorts based on COVID-19 diagnosis. We evaluate the effectiveness of two contact-targeted interventions. In the resident cohorting intervention, recovered residents are moved back from the COVID (infected) cohort to the non-COVID (susceptible/uninfected) cohort. In the immunity-based staffing intervention, recovered staff, who we assume have protective immunity, are assigned to work in the non-COVID cohort, while susceptible staff work in the COVID cohort and are assumed to have high levels of protection from personal protective equipment. These interventions aim to reduce the fraction of people’s contacts that are presumed susceptible (and therefore potentially infected) and replace them with recovered (immune) contacts. We further evaluate two types of screening tests conducted with varying frequency: 1) rapid antigen testing and 2) PCR testing.

**Results:** The frequency and type of testing has a larger impact on the size of outbreaks than the cohorting and staffing interventions. The most effective testing strategy modeled is daily antigen testing. Under all screening testing strategies, the resident cohorting intervention and the immunity-based staffing intervention reduce the final size of the outbreak among residents, with the latter reducing it more. The efficacy of these interventions among staff varies by testing strategy and outbreak size.

**Conclusions:** Increasing the frequency of screening testing of all residents and staff, or even staff alone, in nursing homes has the potential to greatly reduce outbreaks in this vulnerable setting. Immunity-based staffing can further reduce spread at little or no additional cost and becomes particularly important when daily testing is not feasible.

## Introduction/background

In the United States, nursing homes and other long term care facilities have been disproportionately affected by the COVID-19 pandemic.^1,2^ As of September 2020, 40% of COVID deaths nationwide are linked to nursing homes.^3^ Nursing home residents are particularly high-risk for severe symptoms and mortality due to older age and high prevalence of underlying medical conditions that increase risk. In addition, nursing homes have a high risk of transmission because residents live together in close quarters and have frequent close contact with staff.^4^ Furthermore, many facilities suffer from understaffing, which may lead to even higher contact rates between staff and residents, and larger COVID outbreaks.^5^

The ability to develop evidence-based responses to this crisis has been constrained by limited data and a shortage of research in these facilities prior to the COVID-19 pandemic. So far, infection control in many nursing homes has relied on moving infected residents into new rooms in a “COVID cohort,” isolated from other residents, as recommended.^5,6^ Long-term separation after recovery keeps the non-COVID cohort at or near 100% susceptible, and thus has the potential unintended effect of preventing development of institutional herd immunity that could slow outbreaks. Additionally, there is relatively little evidence about the potential of strategies that focus on staffing based on infection histories.

Although guidelines require testing in nursing homes,^6^ the frequency and type of testing varies. In many cases, both cohorting efforts and testing strategies have failed to control outbreaks, likely due in part to identification of infected individuals after transmission has already occured.

A primary driver of these outbreaks is pre-symptomatic and asymptomatic transmission, the latter of which occurs in an estimated 40% of infections.^7,8^ Infected individuals who do develop symptoms become infectious either at or before the time of symptom onset. For this reason, public health screening testing (i.e. testing non-symptomatic individuals) is expected to be an important intervention to control transmission. In addition, control measures that reduce the frequency of contacts among those who are susceptible and those infected but not yet identified could reduce transmission by those who are infectious without symptoms. Due to a lack of resources, it is critical to identify interventions that will most effectively control infection in nursing homes at minimal cost.

Here, we use an agent-based model designed specifically to represent nursing homes in order to examine how a range of interventions may impact COVID-19 outbreaks in this setting. We focus on two contact-targeted interventions to group staff and residents based on their infection and immunity status and examine testing interventions that reflect currently available virological tests. Our proposed strategies involve co-locating susceptible nursing home residents with people – staff and residents – who have recovered and are assumed to be immune, and show that screening with rapid antigen testing outperforms most PCR-based testing strategies.

## Methods

### Model structure

We developed a stochastic, agent-based Susceptible–Exposed– Infectious(Asymptomatic/Symptomatic)–Recovered (SEIR) model to examine SARS-CoV-2 transmission in nursing homes. The model uses a cohorted framework, wherein residents who become infected with SARS-CoV-2 are separated into a distinct population after either showing symptoms or testing positive, where they have no interactions with susceptible (or undiagnosed) residents (Figure 1). Infected residents may be asymptomatic, in which case they can only be identified through testing, or symptomatic, in which case they are identified either by symptom onset or testing, whichever occurs first. Due to continued demands on nursing home capacity throughout the pandemic, we assume that new residents continue to move into the facility to replace those who have died or been discharged, keeping capacity at 100%, and leading to a constant inflow of susceptible residents.

**Figure 1.**
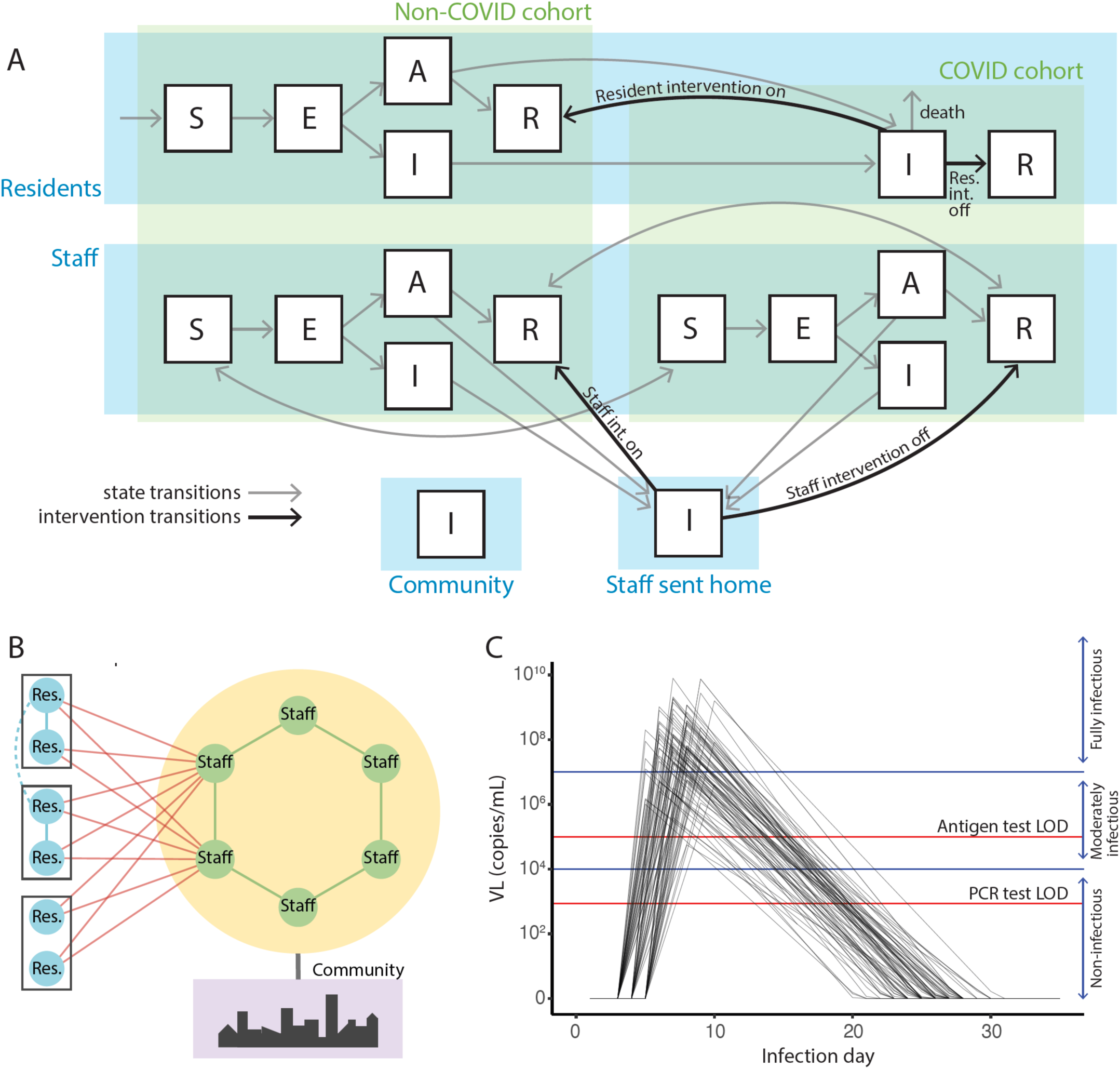
A) Agents move through a stochastic *Susceptible–Exposed– Infectious(Asymptomatic/Symptomatic)–Recovered (SEIR)* compartmental model. The model uses a cohorted framework wherein nursing home residents who become infected with SARS-CoV-2 are moved out of the non-COVID cohort and separated into a distinct COVID cohort after showing symptoms or testing positive. Black lines denote cohorting interventions and grey lines show transitions between infection states. B) Simplified representation of the contact structure within the model framework. There are two residents per room, and each interacts with six staff per day. Staff interact with two other staff each day and also have a daily risk of infection from the community. Sensitivity analyses add additional limited contacts between residents (dashed line, see Figure S7). C) Following a three to five day latent period, viral load is modeled to increase rapidly for two to five^9^ days before peaking. Viral load then immediately begins to decline. Infectiousness of individuals is based on viral load and is modeled categorically: not infectious, moderately infectious, and fully infectious. LOD = limit of detection.

Staffing remains constant, but staff are split between COVID and non-COVID cohorts in proportion to residents. Staff who become infected are sent home to recover before coming back to work, and are replaced by temporary workers while they recover. Daily contact rates are modeled between residents, staff, and between the two populations (i.e. staff-resident contacts). The manner and frequency of potentially infectious contacts between residents and staff are based on contact rates in a Massachusetts network of nursing homes.^10^ In addition, staff have a constant daily risk of infection from the community. The probability of infection from the community is set to approximate different levels of community prevalence (Table S1).

### Viral load and infectiousness

For each individual, viral load follows a tent-function approximating current data on viral shedding as described in the literature (Figure 1C).^9,11^ Following a three to five day latent period, viral load increases rapidly for two to five days before peaking. Viral load then immediately declines. For symptomatic individuals, onset of symptoms occurs after two days of increasing viral load, at which point they can be identified, resulting in an incubation period of 5-7 days.^12,13^ We assume the same viral load trajectories for asymptomatic and symptomatic infections.^14^ The peak viral load varies between individuals, and is drawn from a normal distribution on the log scale around the estimated mean peak viral load.^15^ The duration of detectable viral load varies between individuals and is normally distributed. Viral load can be detected after individuals are no longer infectious.^16^ We base infectiousness categorically on viral load: not infectious, moderately infectious, and fully infectious (Table S1 and Figure 1C). We set the probability of infection given an infectious contact so as to get a plausible R0 in the absence of interventions in our simulations (Table S1).^17^

### Interventions

Using this framework, we evaluate interventions that could be implemented in individual nursing homes: two targeting contact patterns (between residents and/or staff), and variations on two primary testing interventions—PCR and rapid antigen tests (Table 1). Testing interventions begin immediately, while cohorting interventions start three weeks into the outbreak. We assume that at baseline, under current guidelines,^6^ residents who are identified as infected move into isolation in a separate COVID cohort; based on information from a Massachusetts network of nursing home facilities,^10^ we further assume that these residents are not moved back into the non-COVID cohort once they have recovered. Under our resident cohorting intervention, recovered residents are instead moved back to the non-COVID cohort after they have recovered and are prioritized as roommates for new residents, who are presumed to be susceptible. The immunity-based staffing intervention prioritizes placing available recovered staff, who we assume are immune, in the non-COVID cohort, leaving susceptible staff to work in the COVID cohort. Importantly, we assume staff working in the COVID cohort are provided adequate personal protective equipment (PPE) that reduces infection risk. Because we assume that the contacts in nursing homes are necessary, our interventions do not reduce contacts, but rather change with whom those contacts occur. Whenever possible, these interventions reduce the fraction of people’s contacts that are presumed susceptible (and therefore potentially infected) and replace them with recovered (immune) contacts.

**Table 1.** Interventions Summary

| **Contact-targeted interventions** |  |  |  |
| --- | --- | --- | --- |
| Resident: Return recovered residents to rooms in the non-COVID cohort |  |  |  |
| Immunity-based staffing: Place recovered staff in the non-COVID cohort and susceptible staff in the COVID cohort with PPE |  |  |  |
| **Testing interventions (Main)** |  |  |  |
| **Test** | **Staff Frequency** | **Resident Frequency** | **Features\*** |
| None | N/A | N/A | Limit of detection = $10^5$<br>Turnaround time = same day |
| Antigen | Weekly | Weekly |  |
| Antigen | Weekly | N/A |  |
| Antigen | 2.3x/Week | 2.3x/Week |  |
| Antigen | 2.3x/Week | N/A |  |
| Antigen | Daily | Daily |  |
| Antigen | Daily | N/A |  |
| PCR | Weekly | Weekly | Limit of detection = $10^3$<br>Turnaround time = 2 days |
| PCR | 2.3x/Week | 2.3x/Week |  |
| PCR | Daily | Daily |  |

| Testing interventions (Supplement) |  |  |  |
| --- | --- | --- | --- |
| Mix | Daily antigen | Weekly PCR |  |
| Antigen | 3.5x/Week | 3.5x/Week |  |
| Antigen | 3.5x/Week | N/A |  |
| Antigen | Daily | Daily | Limit of detection = $10^7$ |
| PCR | Weekly | Weekly | Turnaround time = 7 days |
| PCR | Weekly | Weekly | Turnaround time = 1 days |

\*Unless otherwise stated, assume:
- PCR test limit of detection = $10^3$ and turnaround time = 2 days - Antigen test limit of detection = $10^5$ and turnaround time = same day

In the testing interventions, we evaluate two types of screening tests: 1) rapid antigen testing, and 2) PCR testing. We evaluate testing at three different frequencies, targeted at either staff only or at the entire nursing home (Table 1). Based on the current parameters of these tests, antigen testing is assumed to have a higher limit of detection (LOD) and consequently lower sensitivity than PCR (10^5 vs. 10^3 copies/mL LOD) but returns results immediately, whereas PCR has a two day delay (Tables 1 and S1). Individuals who are waiting on test results are assumed to behave as normal unless they experience symptoms before receiving results, in which case residents are moved to the COVID cohort and staff are sent home. We further explore variations in the sensitivity of the antigen tests and the turnaround time of the PCR tests (Tables 1 and S1). To compare the efficacy of these strategies, we measure cumulative infections after six months and calculate the mean of 100 stochastic simulations under each combination of contact-targeted and testing interventions.

### Sensitivity analyses

We conduct sensitivity analyses of our assumptions about epidemic dynamics and viral kinetics, varying the prevalence of COVID transmission in the community, the efficacy of PPE, and the relationship between viral load and infectiousness (Table S1). Additionally, we relax the assumption of perfect specificity of antigen testing among staff; in these simulations, we assume a two day delay in confirmatory testing, during which time staff are sent home and are replaced by temporary staff. To examine the impact of our assumptions about nursing homes, we conduct sensitivity analyses around the ratio of staff to residents and resulting contact patterns, as well as increased contact rates between residents.

## Results

We find that the contact-targeted interventions primarily impact cumulative incidence in residents. Because we assume a constant rate of introduction from the community, the change in staff infections within the nursing home due to these interventions is often compensated for by infections acquired in the community. Under all screening testing strategies, the immunity-based staffing intervention, which involves susceptible staff working with infected residents and recovered staff working with susceptible residents, reduces the cumulative incidence of the epidemic among residents. Among staff, the relative efficacy varies by testing strategy (Figure 2, Table 2), but the average cumulative incidence across the nursing home under this intervention is never substantially increased under our baseline parameters (Table 2). This intervention requires sufficient PPE for staff working with infected residents; in the absence of sufficient PPE, it still protects residents but leads to a larger total outbreak among staff (Figure S1). The resident cohorting intervention reduces the cumulative incidence among residents under all testing scenarios but less so than the staffing intervention. Combining the cohorting and staffing interventions provides little added benefit over the resident intervention alone (Figure S2, Table 2) and in scenarios with larger outbreaks, performs worse than the staff intervention alone. Results of each intervention and testing scenario were qualitatively consistent across all 100 simulations (Figure S3).

**Table 2.**
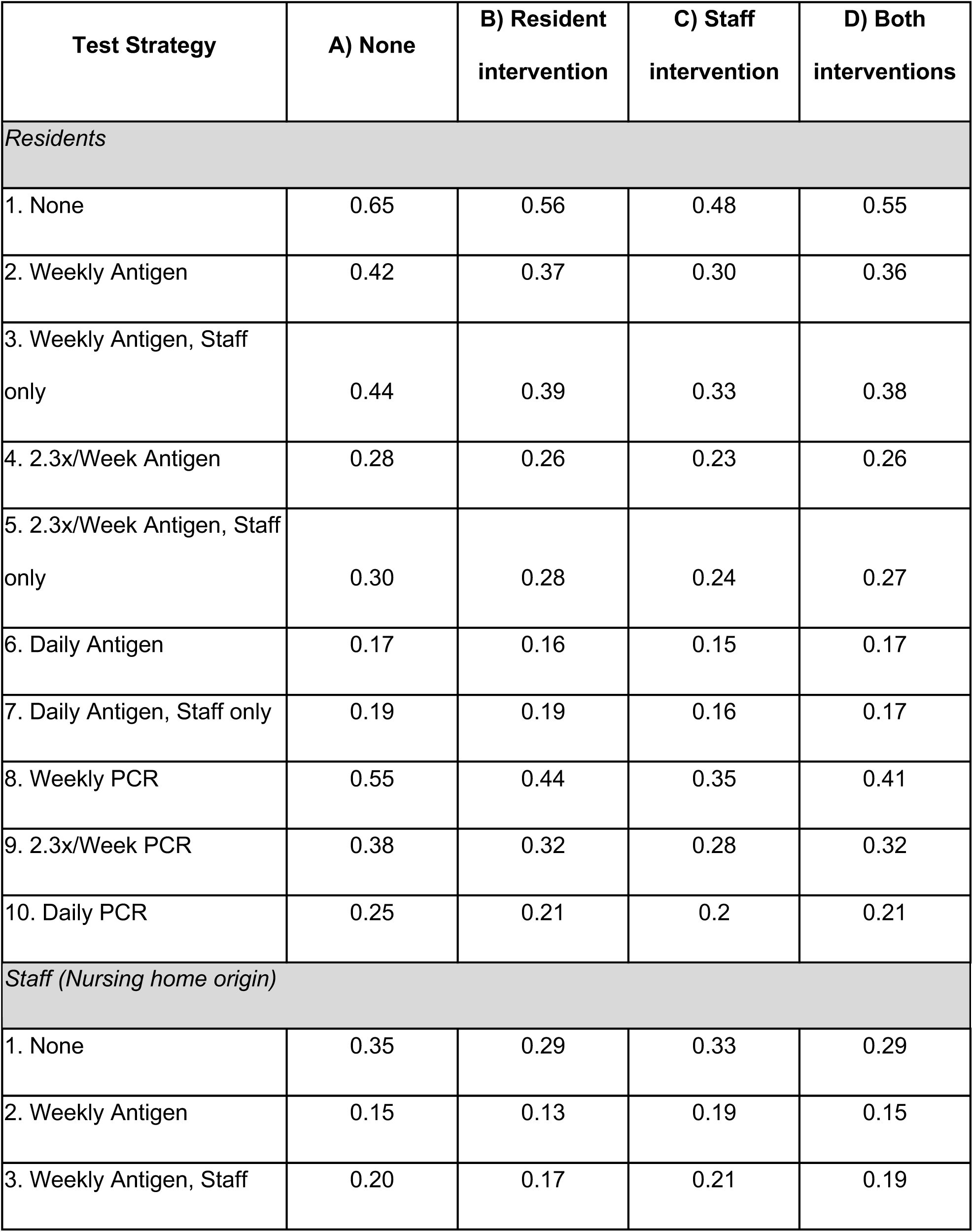
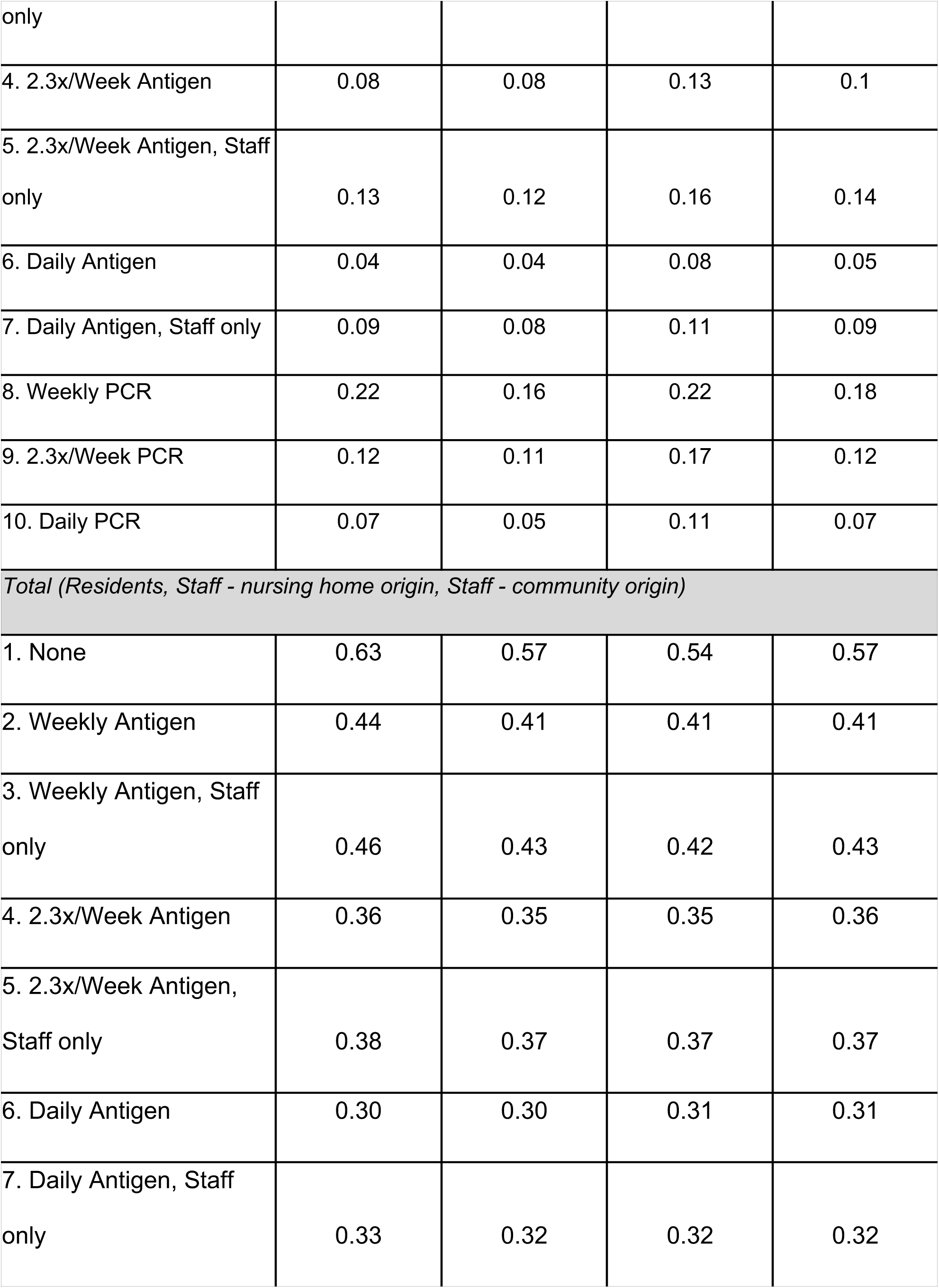
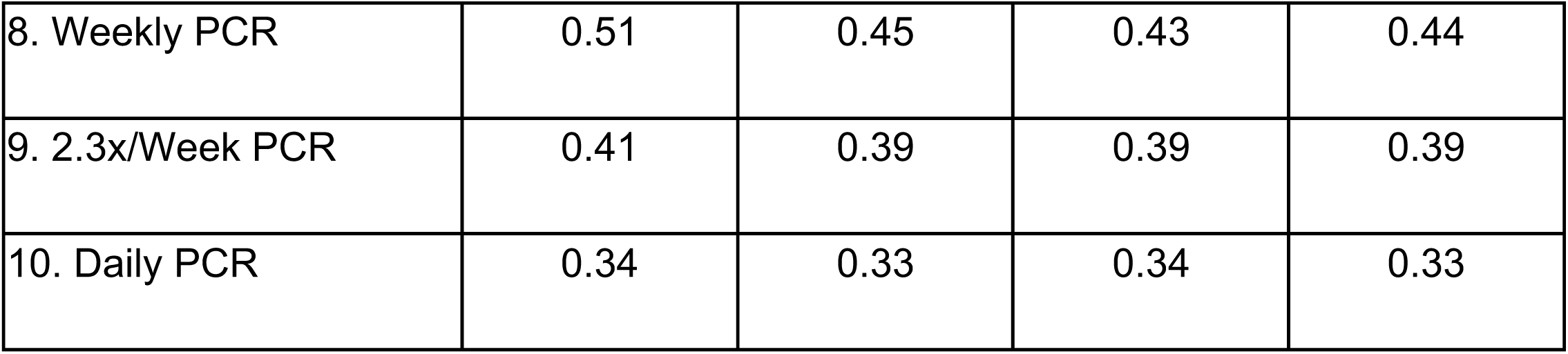
Cumulative incidence

**Figure 2.**
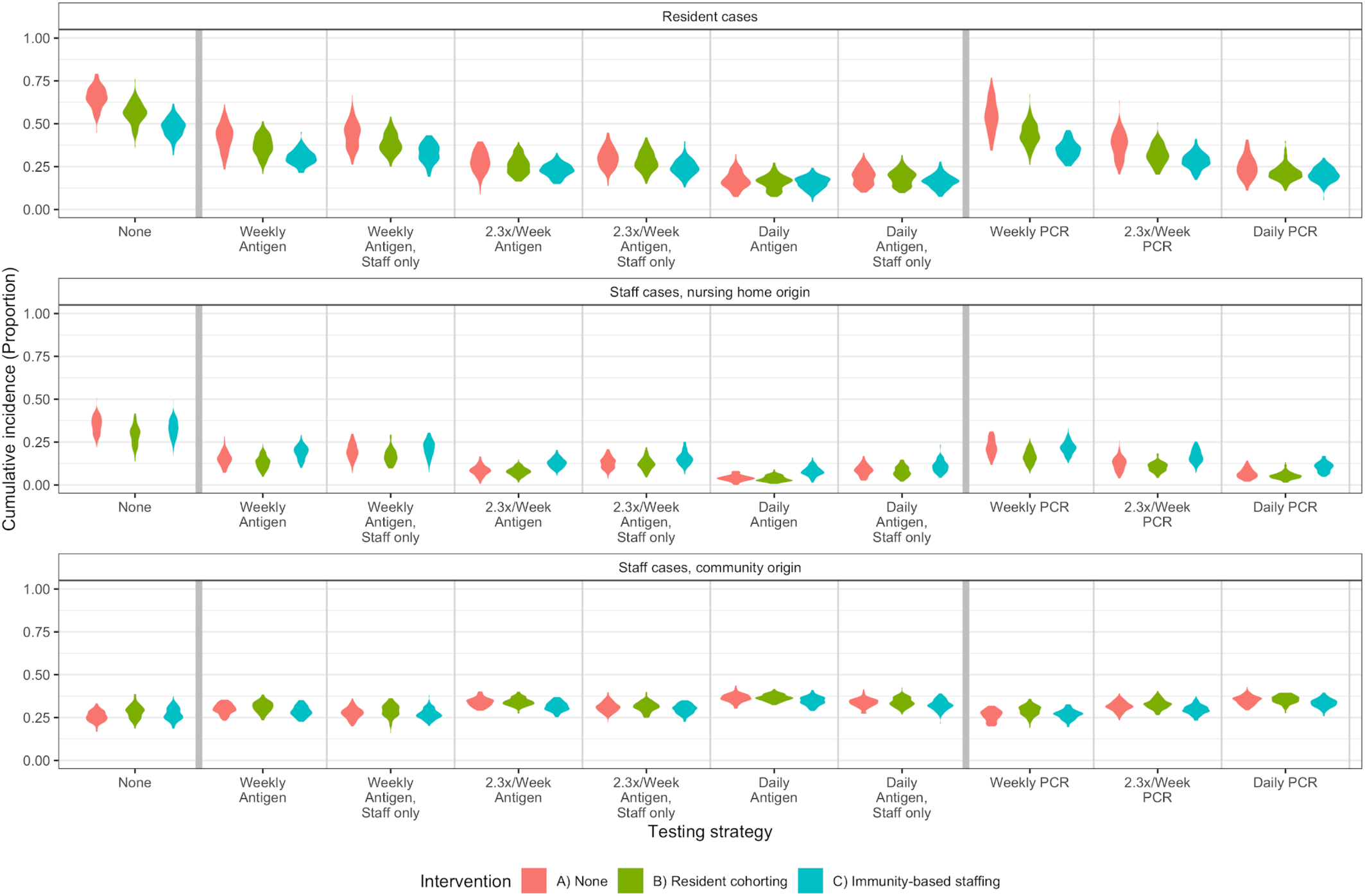
Cumulative incidence at 6 months from first SARS-CoV-2 introduction under different cohorting, staffing, and testing interventions in residents (top panel) and staff (middle and bottom panels). Violin plots show simulation results from 100 stochastic simulations. Cases among staff are split out to distinguish between cases arising in the community and those that are a result of transmission within the nursing home. The staffing intervention reduces the final size of the outbreaks among residents more than the resident intervention; among staff, the relative efficacy varies by testing strategy. Because we assume a constant rate of introduction from the community, the change in staff infections within the nursing home due to these contact-targeted interventions is generally compensated for with infections due to ongoing community transmission. Despite lower sensitivity (LOD = 10^5 unless otherwise specified), rapid antigen testing proves more effective than PCR testing (LOD = 10^3) done at the same frequency due to its faster turnaround time. Antigen test results are returned the same day, while PCR tests are assumed to take 2 days. LOD = limit of detection.

The frequency and type of testing had a much larger impact on the size of epidemics than the cohorting and staffing interventions (Figure 2). We examine both frequency and “quality” of testing: for antigen tests we explore different sensitivities and for PCR testing we examine the impact of turnaround time, since these are the most important limitations of each type of test (Figure S2). For each test we model daily, every two day, or weekly testing, either of staff and residents or staff only. We find that the most effective testing strategies are daily antigen testing of everyone and daily antigen testing of staff only. Despite lower sensitivity, these strategies prove more effective than PCR testing done at the same frequency due to the faster turnaround time. However, if antigen tests have very low sensitivity (LOD = 10^7 vs. baseline assumption of 10^5 copies/mL; Figure S2), then daily antigen testing is less effective than daily PCR testing. In the baseline simulations, we assume perfect specificity, for example through the use of rapid orthogonal molecular or antigen based confirmatory tests whenever a rapid antigen test is positive. When we relax this assumption and assume that confirmatory tests have a two day lag, we see no change in the relative efficacy of interventions and testing strategies; however, cumulative incidence is higher (Figure S4). Imperfect specificity increases the number of temporary staff required, especially at greater testing frequencies, which in addition to increasing the opportunities for importations, may also be an additional burden on nursing home operations.

To examine the impact of ongoing issues with testing delays, we compare a scenario with weekly PCR testing with a 7 day turnaround time, rather than our baseline assumption of 2 days. In these simulations, more residents are infected after 6 months than in simulations with no testing at all (Figure S2). This occurs because outbreaks without testing peak more rapidly than those with testing, leading to herd immunity in the facility that protects residents who enter the facility later from getting infected. In contrast, testing with long delays draws out the outbreak, allowing more incoming residents to become infected (Figure 3). Under more effective testing strategies, such as daily antigen testing, outbreaks are effectively prevented and cumulative incidence primarily reflects continued introductions from the community (Figure 3).

**Figure 3.**
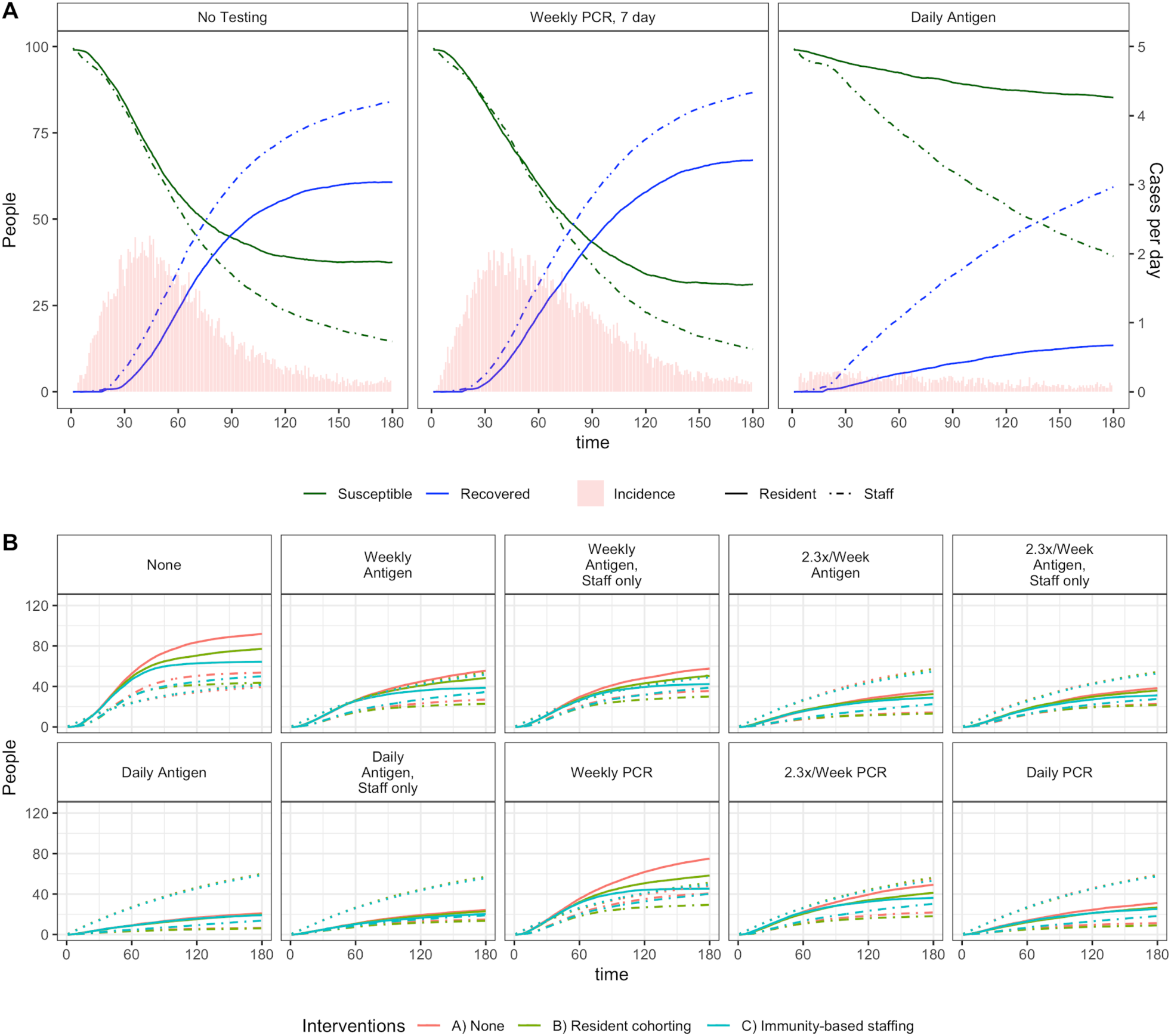
A) Mean outbreak dynamics in the absence of cohorting and staffing interventions. Mean daily incidence is shown on the right axis. Left panel: no testing—outbreak occurs more rapidly. Center: weekly PCR, with a 7 day delay, results in slower outbreak and larger cumulative incidence over six months. Right: daily antigen testing prevents a recognizable outbreak curve, and results in the smallest cumulative incidence of all testing measures. Due to constant risk of infection from the community, we see continued introductions and spread within the nursing home. B) Mean cumulative incidence proportion over 6 months of simulation in each baseline testing scenario. Solid lines indicate cases in residents; dotted lines indicate cases in staff that were introduced from the community, and dashed lines indicate cases in staff that occurred inside the nursing home.

Community prevalence has an important impact on outbreak size and efficacy of interventions (Figure 4, Figure S5). When it is lower, intervention efficacy follows the same general trends but is reduced for contact-targeted interventions, particularly when combined with more effective testing strategies. In addition, reducing the frequency of testing has a smaller effect on final size. In this setting, weekly antigen testing is approximately as effective as daily antigen testing, and 2.3x weekly PCR is not clearly different from daily PCR—suggesting that in settings with lower prevalence, lower frequency testing may suffice. At higher community prevalence, outbreaks (i.e. any onward transmission) always occur due to constant seeding but the size of the outbreak depends on testing frequency and interventions (Figure 4, Figure S6). At lower prevalences, the probability of an outbreak depends on the testing frequency, but once an outbreak occurs testing frequency has less impact on outbreak size.

**Figure 4.**
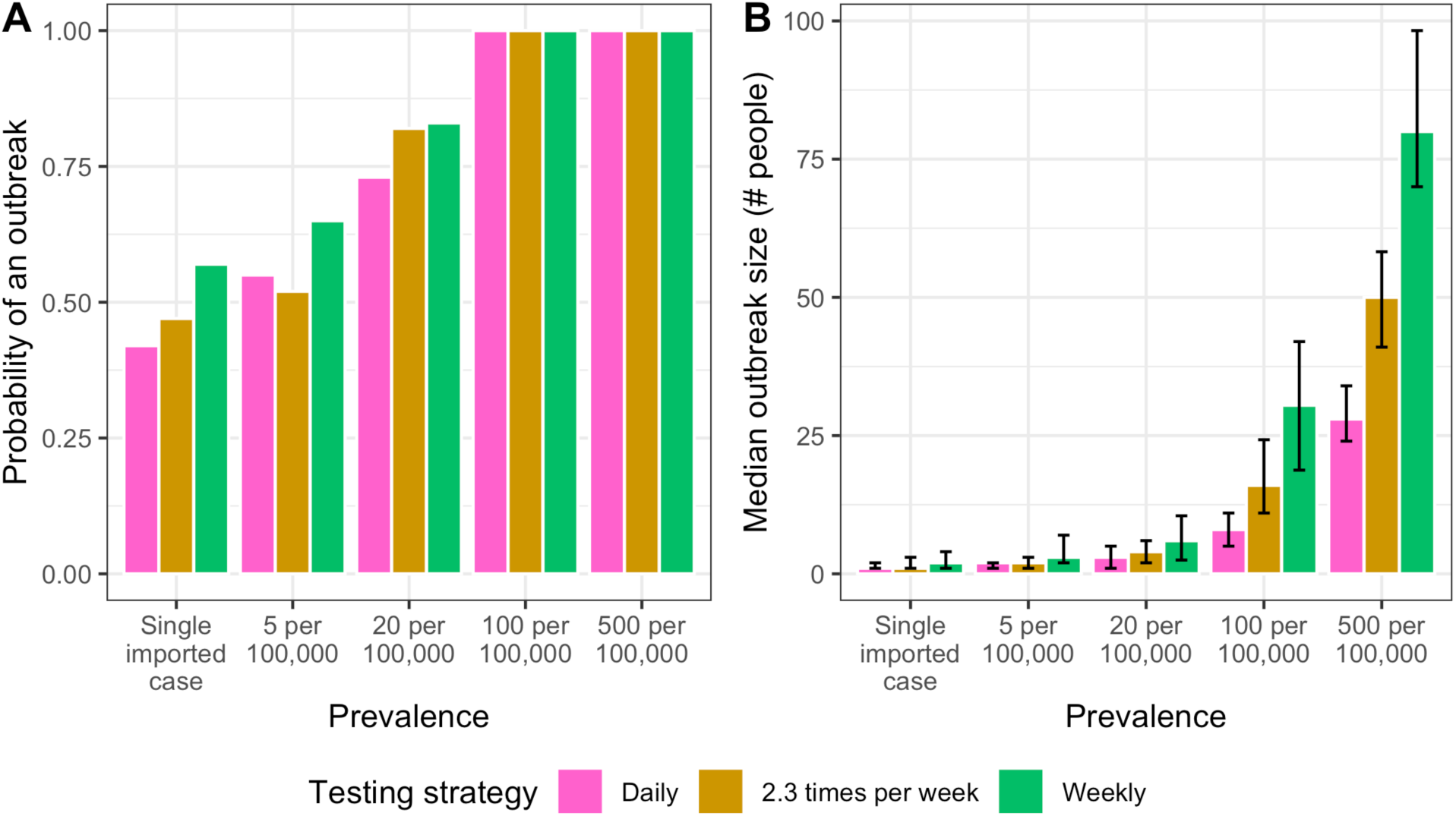
A) Probability of an outbreak, defined by a single onward transmission event, across four different community prevalence levels. The far left column shows results when there was a single initial case but no additional community introductions (i.e. no community prevalence). B) Median outbreak size (# people) given that an outbreak (i.e. ≥1 onward transmission) did occur, with interquartile ranges.

Increasing contact rates, either through non-roommate interactions between residents (Figure S7) or decreasing the staff to resident ratio and therefore increasing the number of resident contacts for each staff member (Figure S8), leads to larger outbreaks. Although the overall cumulative incidence is sensitive to these parameters, the relative efficacy of testing and cohorting and staffing interventions does not change.

## Discussion

Increasing the frequency of screening testing of all residents and staff, or even staff alone, in nursing homes has the potential to greatly reduce outbreaks in this vulnerable setting. Our results provide further support to other models’ findings that faster test results should be prioritized over high test sensitivity,^11,18^ even in small congregate settings like nursing homes. While rapid antigen testing generally performs better than PCR testing—which is more sensitive, but has a longer turnaround and may be prohibitively expensive—we do see that when the limit of detection is very high for antigen testing, this does not hold. This suggests that there may be a threshold effect at which point low sensitivity results in ineffective antigen testing.

The staffing intervention that we propose here can further reduce spread among residents at little or no additional cost and becomes particularly important when daily testing is not feasible. It may have the additional advantage of being more straightforward to implement than an intervention that involves moving residents between rooms or floors, as in the cohorting intervention. The staffing intervention works by reducing the risk that staff could infect susceptible residents by prioritizing recovered staff to work with them, lowering the effective reproduction number in the non-COVID cohort. This intervention hinges on the assumption that recovered staff are protected against reinfection, and access to appropriate PPE is required for it to be feasible. When the resident intervention is combined with the staff intervention, outbreak size is similar to under the resident intervention alone. This may happen because when recovered residents are moved back to the non-COVID cohort, more staff are also moved out of the COVID cohort—increasing the proportion of non-COVID staff who are still susceptible, and diluting the protective effect of the recovered staff in the non-COVID cohort.

Our results suggest that, given limited resources, community prevalence could guide the choice of interventions. When prevalence is lower, contact-targeted interventions have a much smaller effect on cumulative incidence. This is because the fraction of each person’s contacts who are infected is already low, so the marginal benefits of reducing that fraction is small. There is also less gained from increased testing frequency, suggesting that when community prevalence is low and testing resources are limited, nursing homes could consider using weekly testing until an outbreak is detected either inside or outside of the facility. Models of similar settings have also found the frequency of testing required to control outbreaks can be dependent on community prevalence.^19^ In addition, when outbreaks are very large—such as in our sensitivity analysis around the relationship between viral load and infectiousness—we see that the staffing intervention is particularly effective at reducing cumulative incidence among both residents and staff (Figure S9). Expanding this model to identify optimal testing strategies in nursing homes without detected outbreaks^20^ and to explore the timings and thresholds for implementing reactive strategies is an important area for future work.

We have made several simplifying assumptions in our model. We assume transmission only occurs upon contact, and do not take into account the possibility of strictly airborne transmission. We further assume no spillover between the COVID and non-COVID cohorts; in settings with understaffing, this assumption may be violated due to compounding effects of outbreak-related stressors on nursing homes. As with other viruses,^21^ the relationships between viral load and infectiousness and viral load and the antigen test sensitivity are not fully understood. We have simplified this complex process, which can be updated as more data become available. Sensitivity analyses that remove this relationship between viral load and infectiousness result in similar findings regarding relative efficacy of testing strategies (Figure S9). We also assume no differences between symptomatic and asymptomatic individuals and ignore the persistent low detectable viral loads that have been described in SARS-CoV-2 infections:^16^ because we are looking at relatively frequent screening testing strategies, we do not expect dynamics at the end of infection to impact our results. We further assume the sensitivity of the tests only depends on viral dynamics and do not incorporate other factors such as variations in sample quality. Regarding nursing home parameters, we assume that on average residents have long durations of stay; future work should incorporate varying lengths of stay. Finally, we assume that staff have random schedules and do not have repeated resident assignments over time.

We have shown how simple interventions involving testing and staffing can greatly reduce the burden of SARS-CoV-2 in nursing homes. As frequent testing requires additional staff time, the choice of interventions will depend on the resources available. Our model further underscores the importance of rapid screening testing, as well as providing adequate PPE to staff.

## Data Availability

Code is available on github.

https://github.com/rek160/NursingHomeABM

## Acknowledgments

We thank Ron Anglo for helpful discussions and insights into Massachusetts’ nursing homes and Rachel Slayton for helpful discussions and feedback.

## Supplement

**Table S1.**
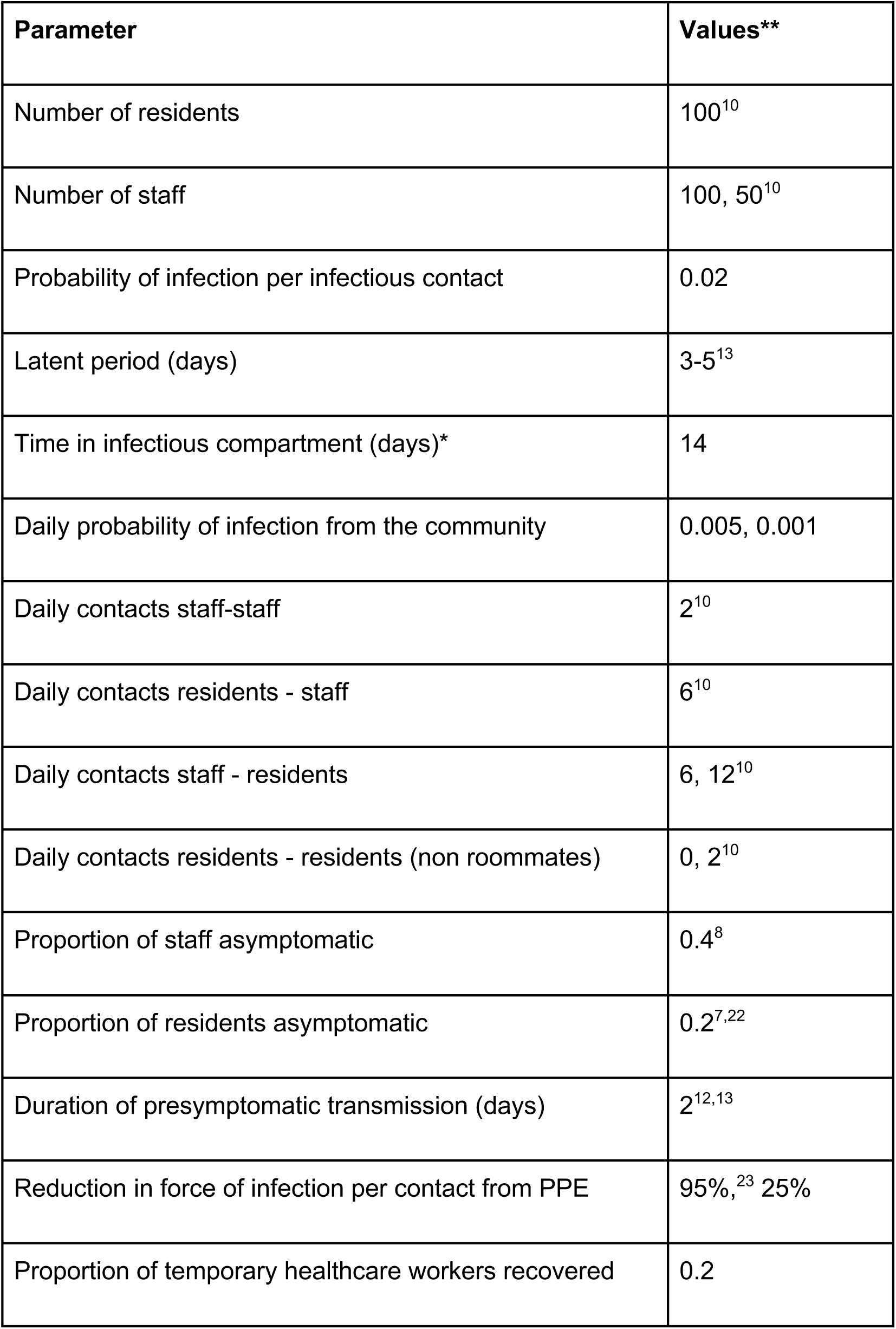
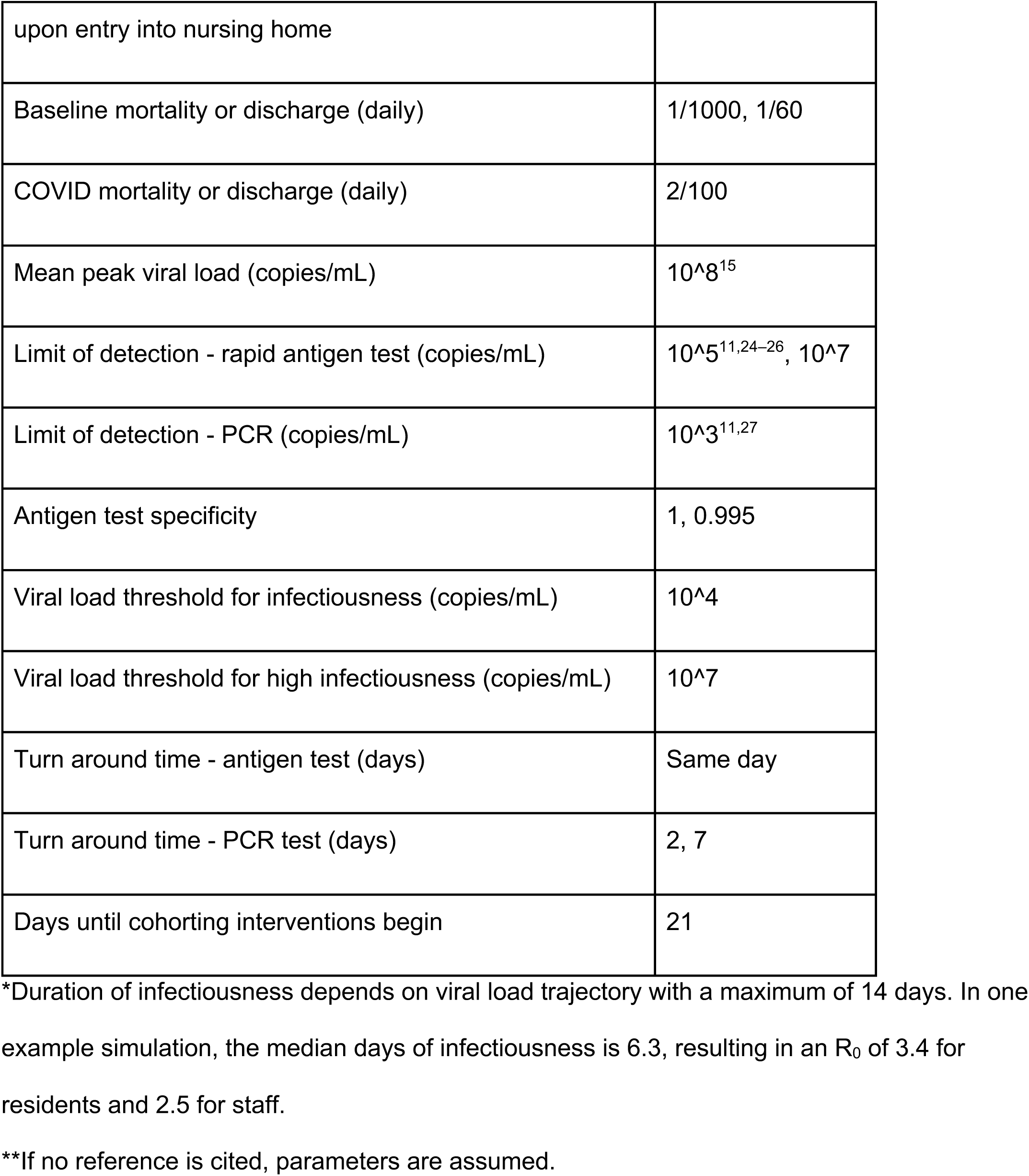
Parameters

**Figure S1.**
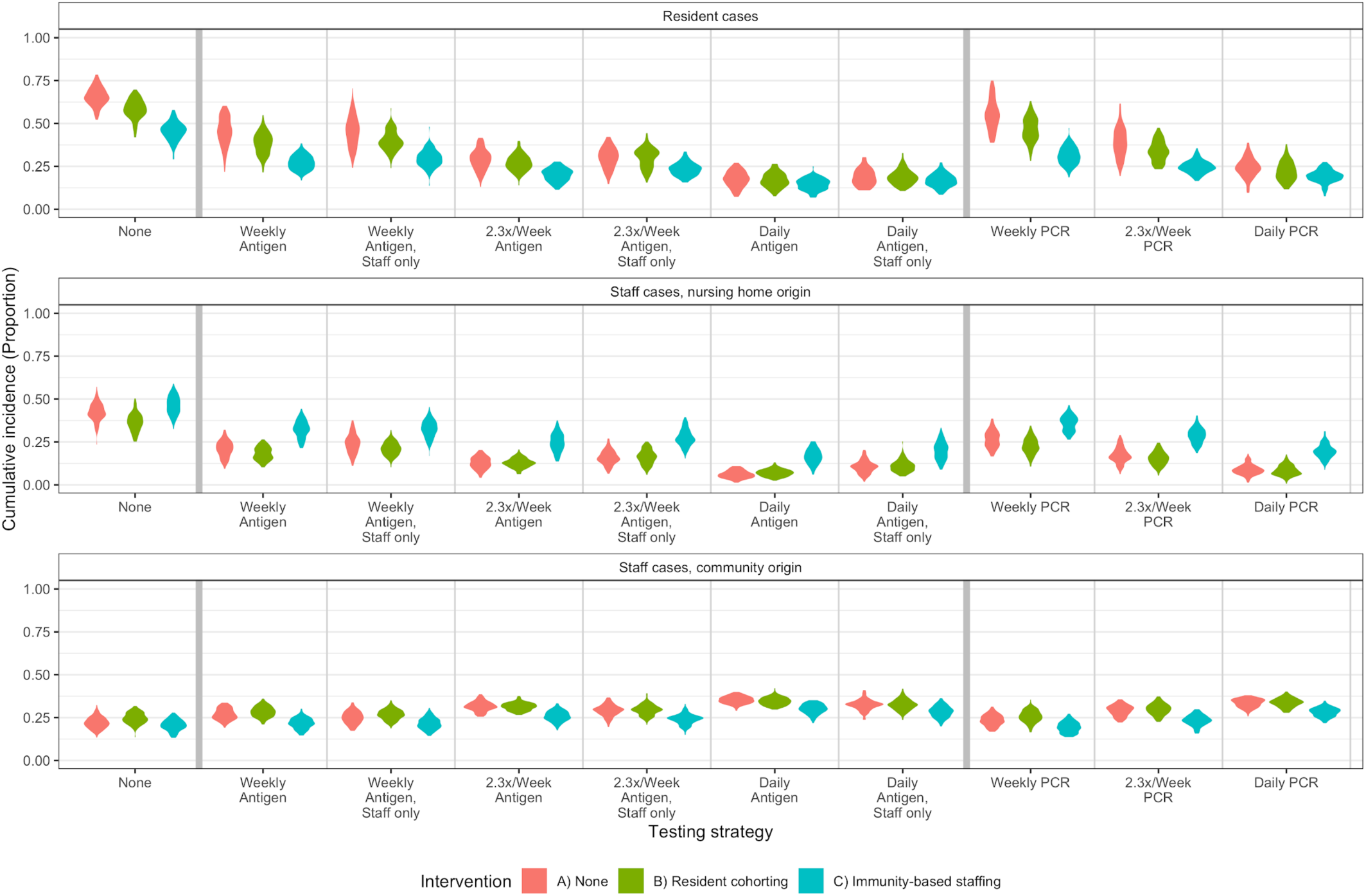
Lower PPE efficacy. When PPE efficacy is lower, intervention and testing efficacy follows the same general trends, but with slightly higher cumulative incidence among staff (nursing home origin).

**Figure S2.**
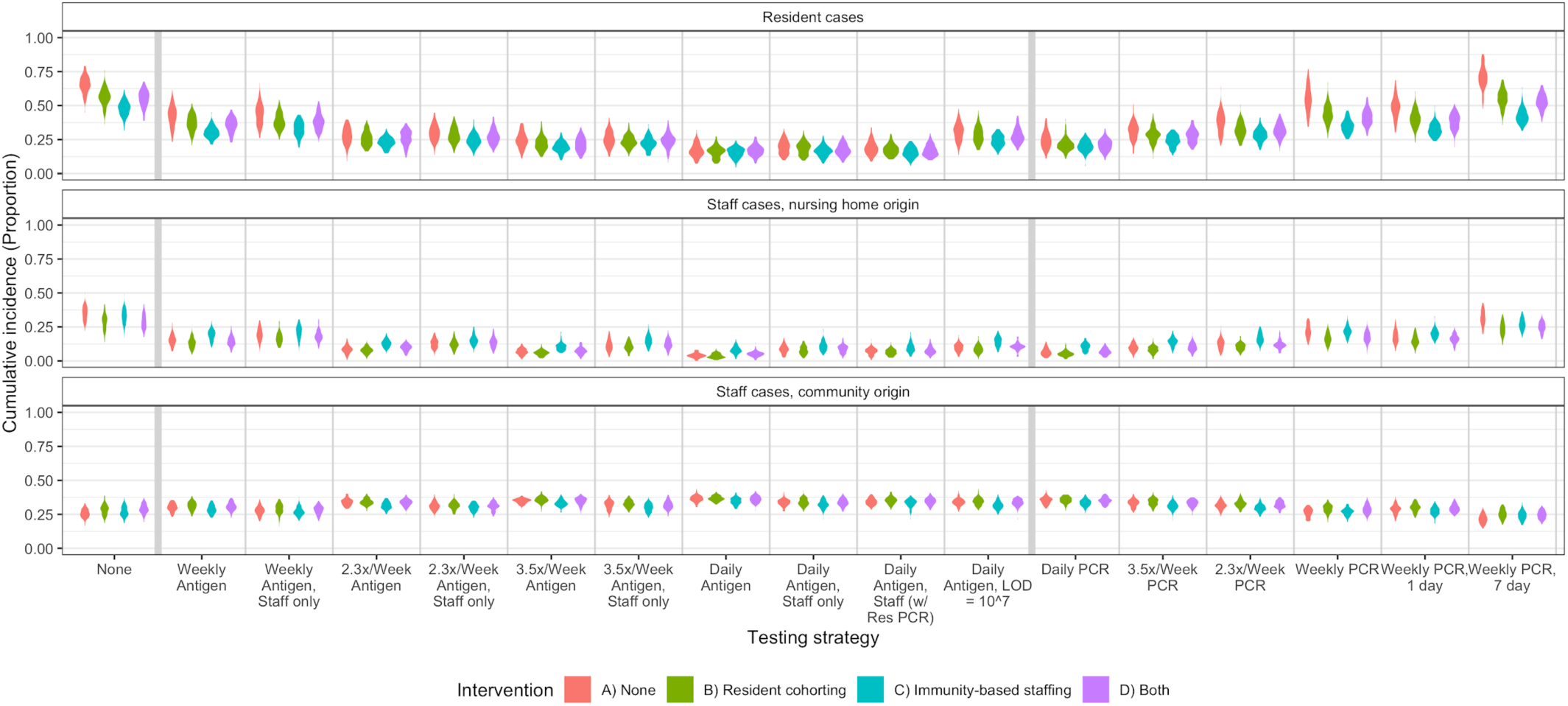
Additional intervention strategies. Cumulative incidence at 6 months from first SARS-CoV-2 introduction under an extended set of testing and intervention strategies. Testing strategies included here but not in the main text (Figure 2) are results for testing every other day (3.5x/week) as well as sensitivity analyses. A combination of the resident cohorting and immunity-based staffing strategies is also shown in purple.

**Figure S3.**
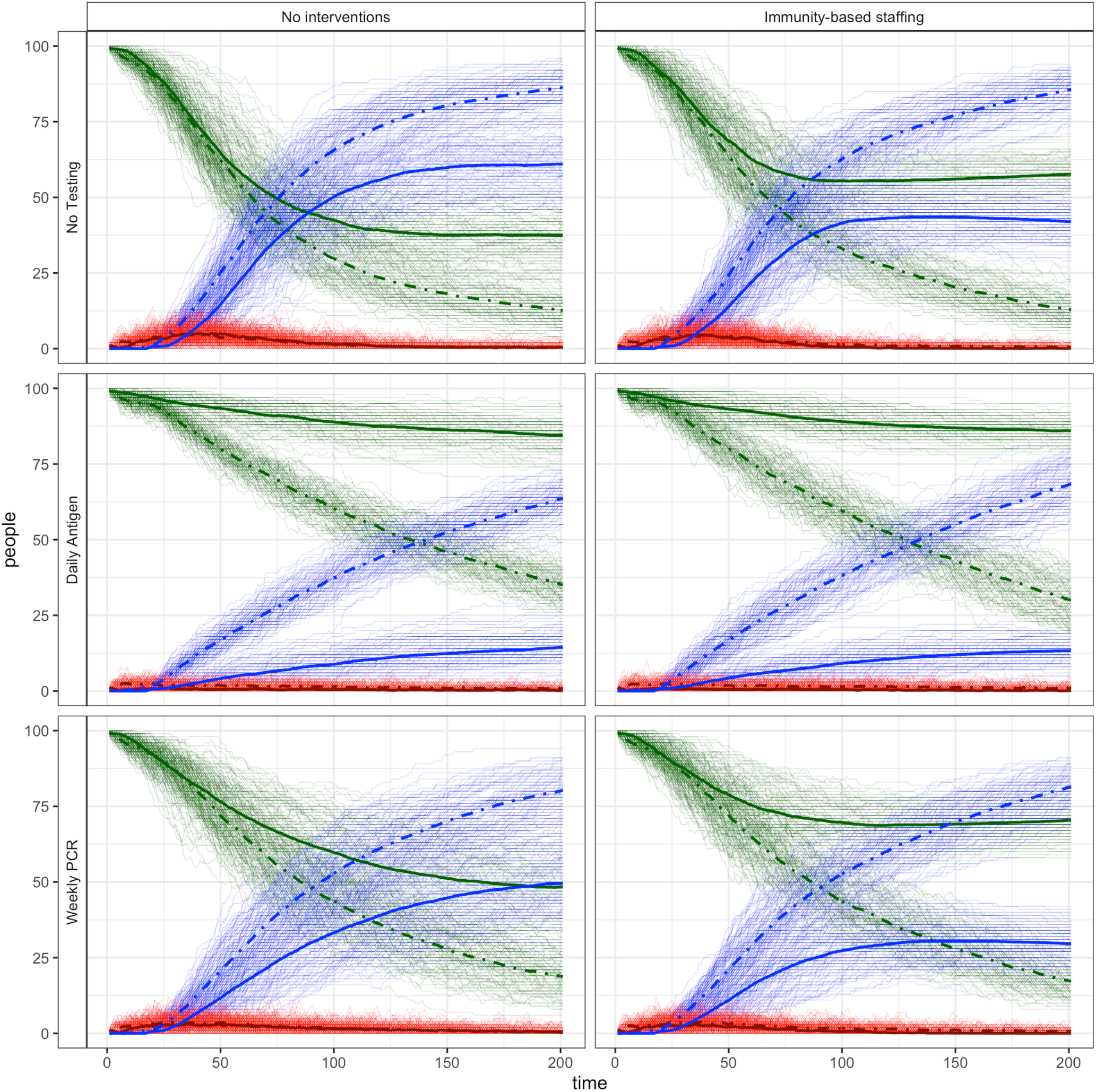
Individual simulations. Plotting individual simulation trajectories show that outbreak dynamics remain relatively consistent across simulations. Solid lines are residents, and dashed lines are staff.

**Figure S4.**
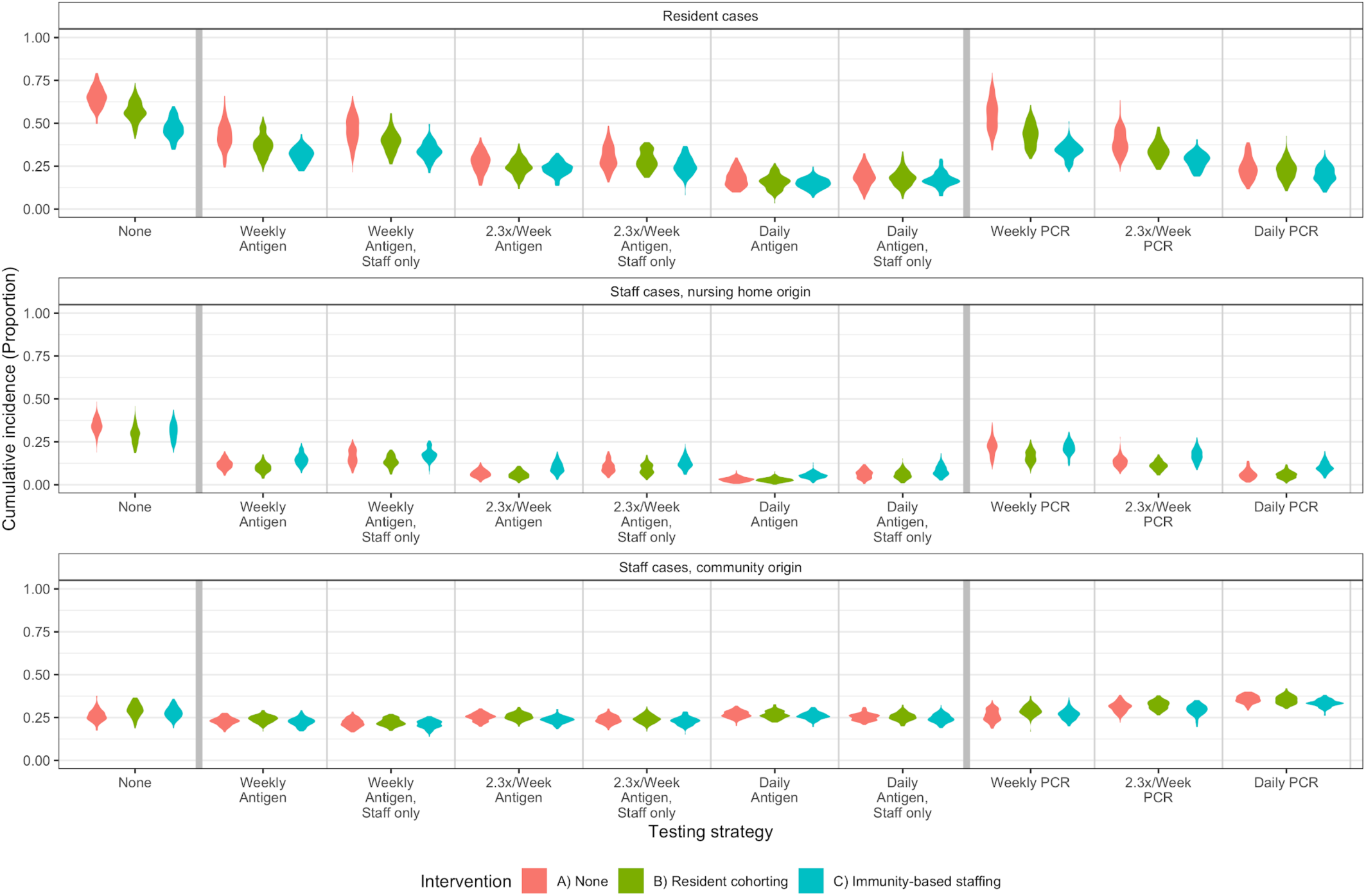
Imperfect specificity. When antigen test specificity is imperfect, intervention and testing efficacy follows the same general trends as when specificity is 100%. However, additional temporary workers are required.

**Figure S5.**
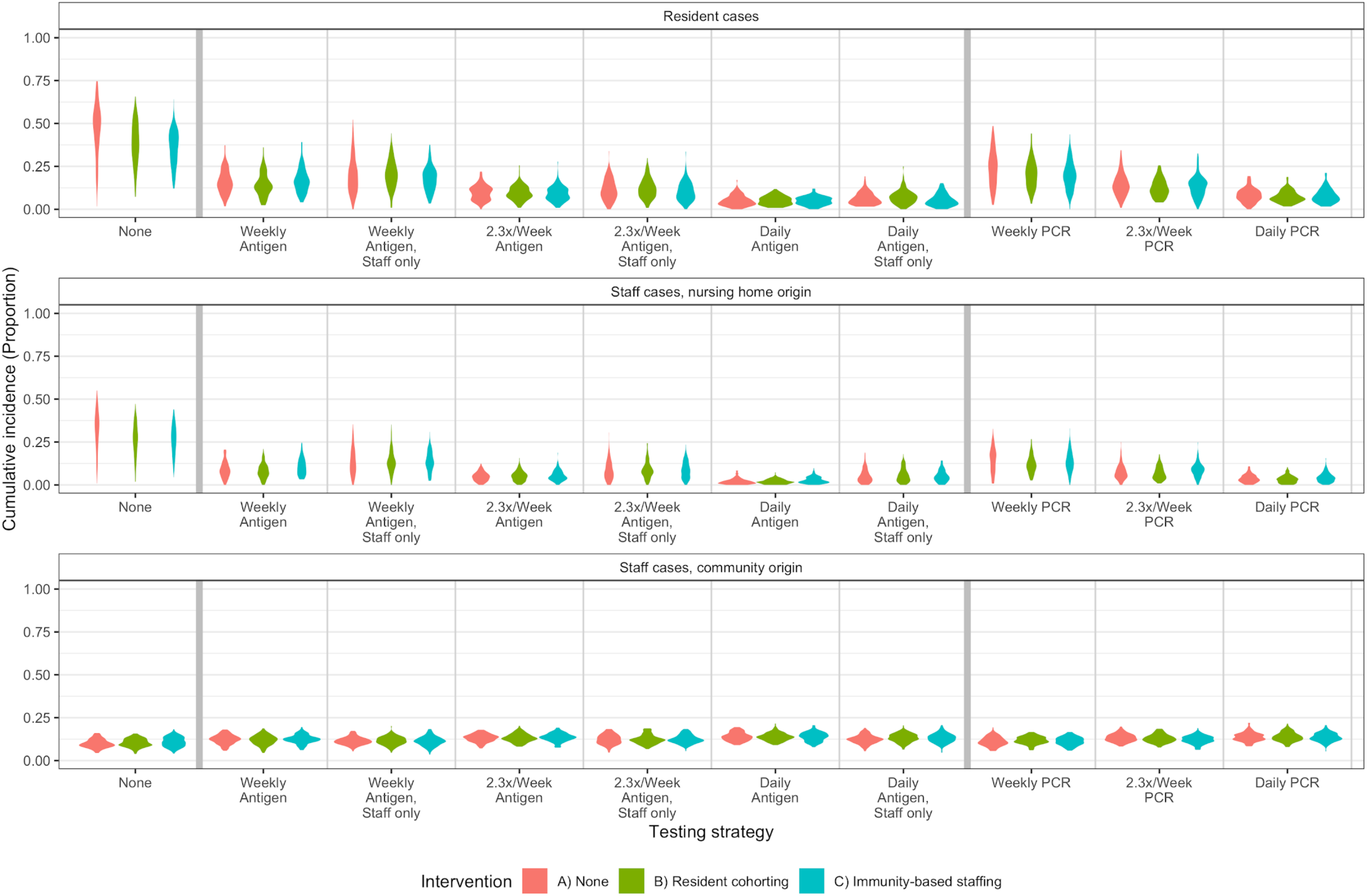
Lower community prevalence. When community prevalence is lower, intervention efficacy follows the same general trends but is reduced for cohorting interventions, particularly for more effective testing strategies. Testing frequency does not impact cumulative incidence as much as at higher prevalences.

**Figure S6.**
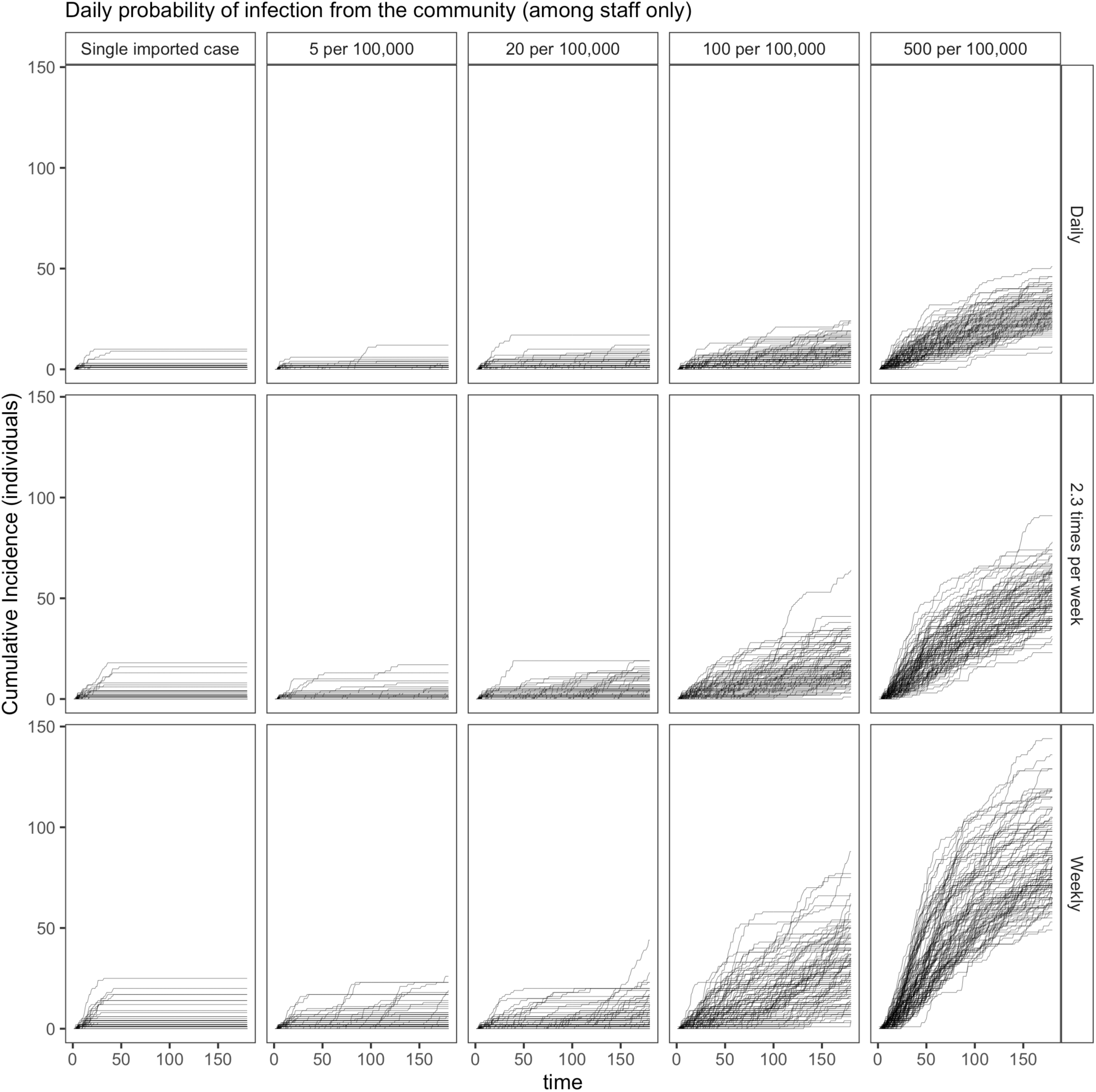
Cumulative incidence by daily probability of infection from the community and testing strategy. Cumulative incidence (100 simulations) by daily probability of infection from the community (columns) and testing strategy (rows). This excludes staff members infected from the community. Although they are less common at lower prevalences (or when seeded with only one case, as in the far left column), larger outbreaks do occur when testing is less frequent.

**Figure S7.**
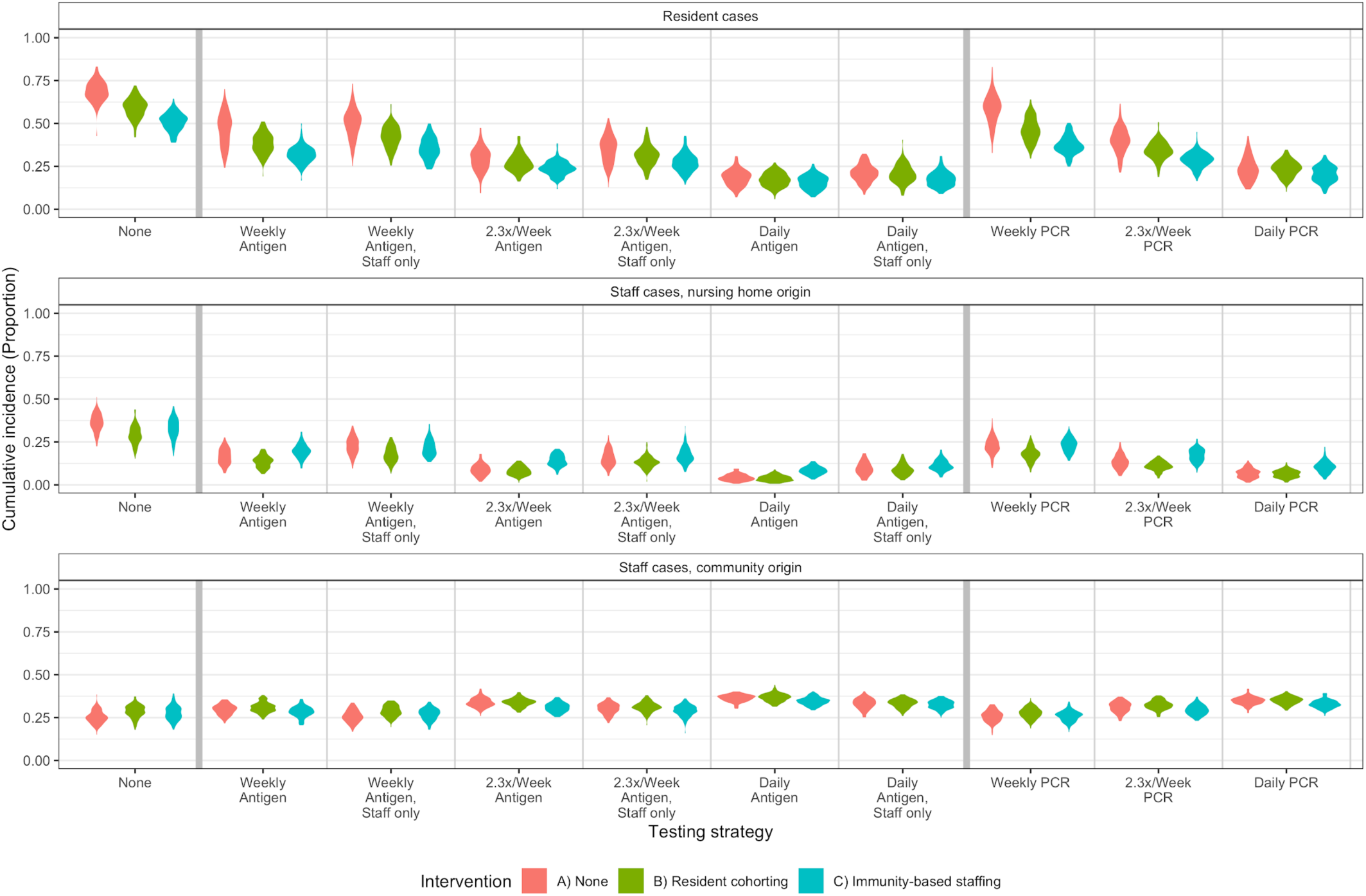
Higher resident-resident contacts. When the number of contacts between residents is increased beyond roommates only, the relative efficacy of interventions and testing does not change.

**Figure S8.**
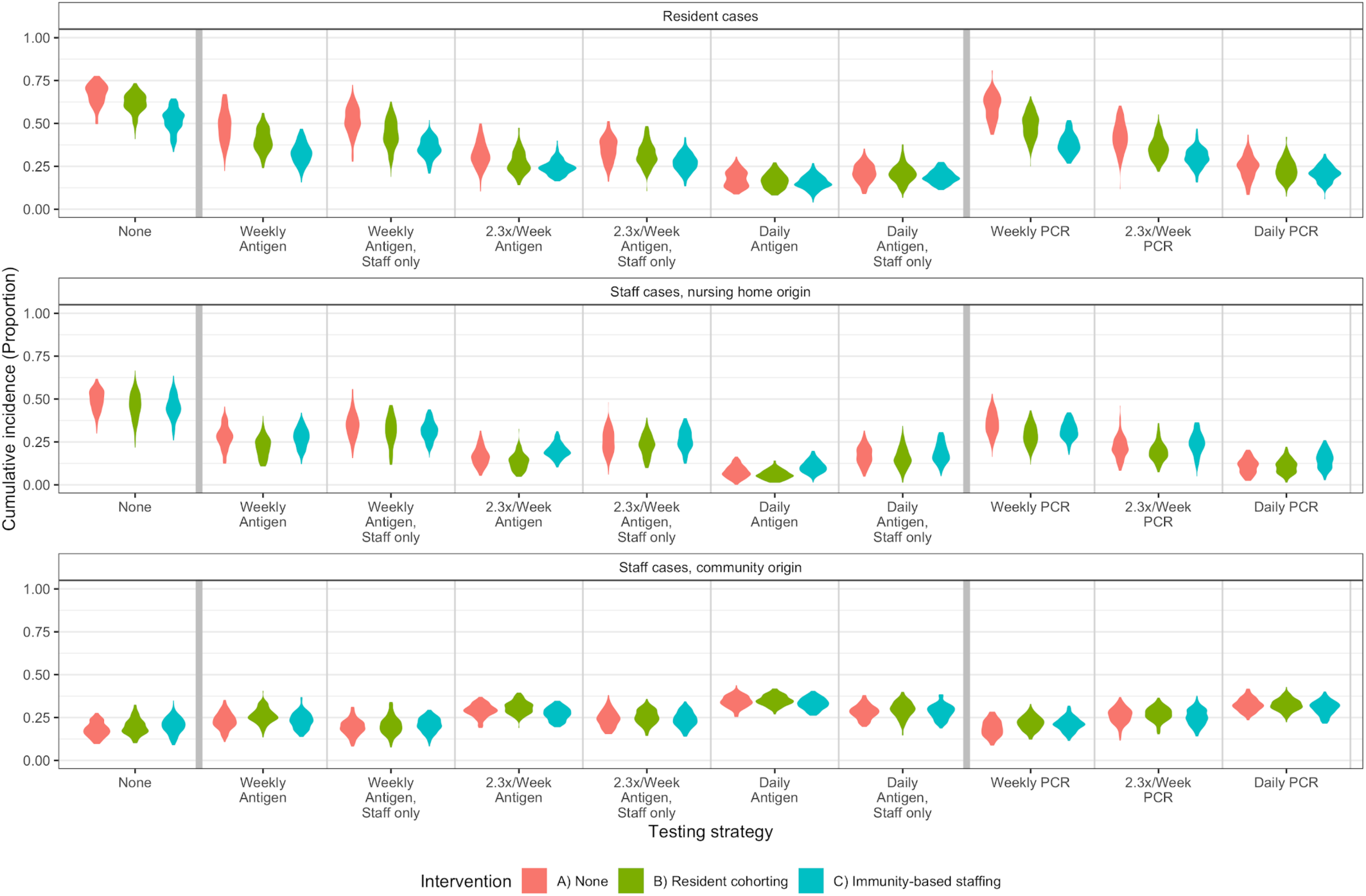
Lower staff-resident ratio. When the ratio of staff to residents is decreased, intervention efficacy does not change.

**Figure S9.**
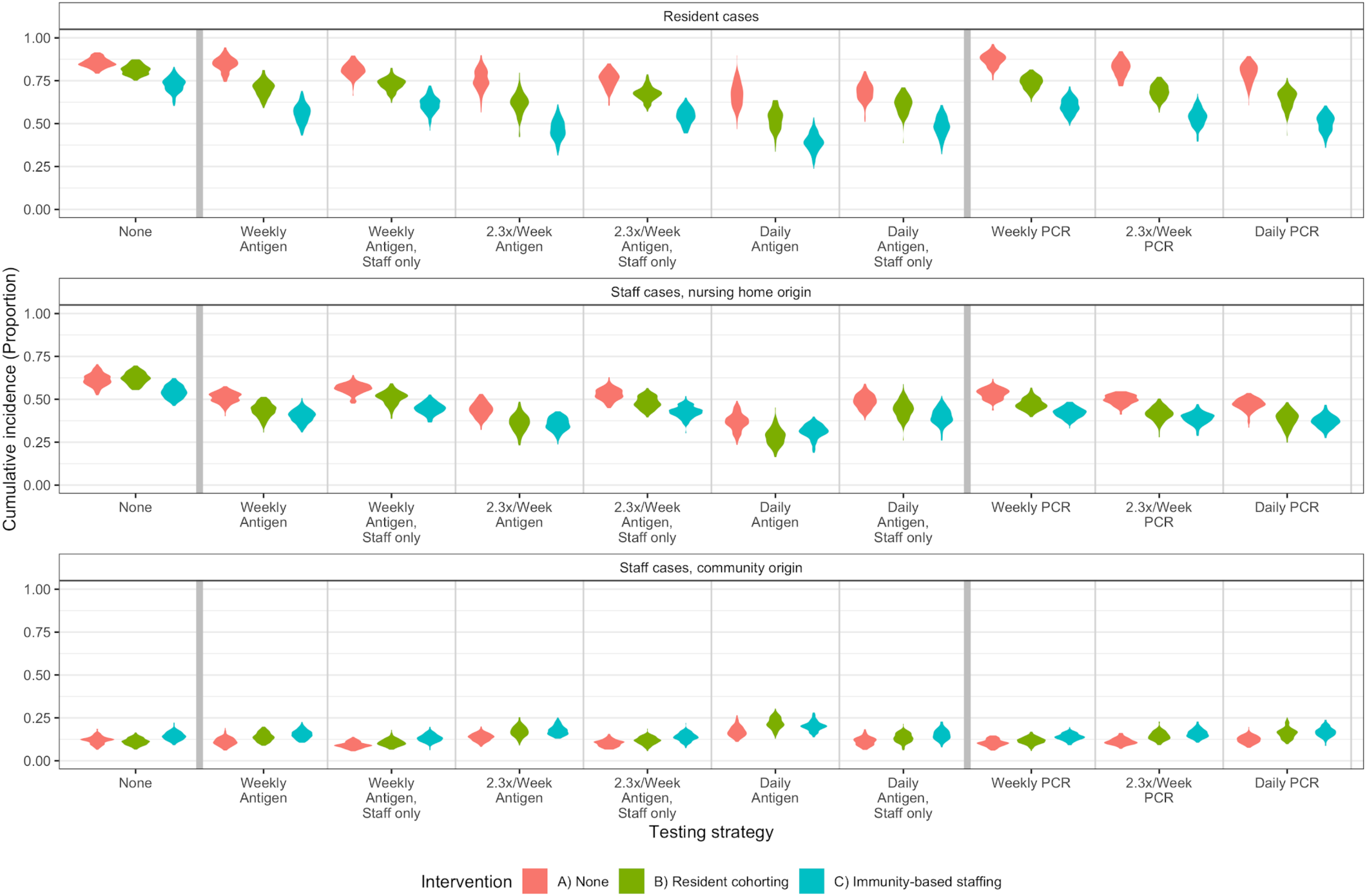
Binary infectiousness, unrelated to VL. When infectiousness begins as soon as VL is detectable, and is not related to VL, we do not see a change in the relative efficacy of our staffing and resident cohorting interventions among residents, although the staffing intervention is more effective in staff than in the baseline simulations. Outbreaks are larger and testing is less effective overall.

